# Antibiotic treatment of asymptomatic urogenital infections in infertile couples is associated with improved sperm quality including sperm apoptosis parameters

**DOI:** 10.64898/2026.09.30.26364433

**Authors:** Jan Novák, Viktor Vik, Milan Drahoš, Alena Langerová, Vojtěch Thon, Zuzana Krátká

## Abstract

**Background:** European Association of Urology (EAU) Guidelines identify urogenital infections as a possible treatable cause of infertility, yet the current recommendation for antibiotic treatment remains weak due to a lack of high-quality evidence. We evaluated the effect of antibiotic treatment of asymptomatic urogenital infections on both conventional and apoptosis-related (flow cytometry sperm apoptosis test, FACS-APO) sperm parameters, as well as on pregnancy outcomes, in infertile couples.

**Methods:** In this retrospective, single-centre study, we analysed 265 couples with an asymptomatic urogenital infection (*Ureaplasma urealyticum*, *Mycoplasma hominis*, *Mycoplasma genitalium*, and *Chlamydia trachomatis*) confirmed in at least one partner by first-void urine testing, who received simultaneous antibiotic treatment. Paired conventional semen analysis and FACS-APO were performed before and 0–9 months after treatment completion. Partner infection concordance, the effect of follow-up timing on treatment response, and pregnancy outcomes within 12 months were also assessed.

**Results:** Sperm concentration, total sperm count, total and progressive motility, and the proportion of viable sperm increased significantly after treatment, while the proportion of late apoptotic / necrotic sperm and calculated sperm DNA fragmentation decreased significantly (all p<0.05); the proportion of early apoptotic sperm remained unchanged, and semen volume did not change. Among 244 couples with complete bilateral microbiological testing, *Ureaplasma urealyticum* was by far the most prevalent pathogen (57.8% of men, 87.7% of women), and female partner positivity given male positivity was 83.0%. The improvement in sperm concentration and total sperm count was most pronounced within 3 months after treatment and diminished, or reversed, beyond 6 months. Pregnancy was achieved in 127 of 265 couples (47.9%; per-protocol rate 53.4%) within 12 months, most commonly following in vitro fertilization (72/127) or spontaneously (21/127).

**Conclusions:** Antibiotic treatment of asymptomatic urogenital infections in infertile couples was associated with significant improvement in conventional semen parameters and a reduction in sperm apoptosis and calculated DNA fragmentation. Given the retrospective, uncontrolled design, these findings should be regarded as hypothesis-generating rather than definitive proof of a causal, fertility-enhancing effect, and warrant confirmation in prospective controlled studies.

## 1. Introduction

European Association of Urology (EAU) Guidelines identify urogenital infections as a possible treatable cause of infertility [1]. The prevalence of these infections and resulting inflammations reaches 6-15% among infertile couples [2]. However, the EAU Guidelines simultaneously refer to a large systematic review by Fode et al. in 2016 [3], which could not have proved any obvious connection between both symptomatic and asymptomatic urogenital infections, incl. those caused by bacteria like *Neisseria gonorrhoeae* or *Chlamydia trachomatis*, on the one hand, and infertility on the other hand owing to a lack of good-quality data. For this reason, no clear recommendation regarding both diagnostic workup and treatment can be drawn. Based on the nature of reproductive medicine (i. e. natural fluctuations of sperm parameters even in healthy individuals, unspecific alterations of sperm parameters, sperm generation time of almost 3 months, idiopathic infertility in 30-50% of infertile males [4], and finally age and more precisely gynaecological finding of the female partner as the most important factor of male infertility), strong evidence-based data are barely achievable.

Patients with symptomatic urogenital infections are being examined by urologists or venereologists for more or less specific complains like dysuria, urgency, urethral discharge or just a discomfort localized to the genital. Detection of the pathogen is followed by an adequate antibiotic treatment to relieve the patient of the symptoms. Whereas infertile couples are typically diagnosed with asymptomatic urogenital infections, which by definition do not cause any complaints. Regarding the impact of asymptomatic urogenital infections – urethritis, epididymitis, prostatitis/prostatovesiculitis – collectively termed as “male accessory gland infections” (MAGIs) [2], there are two completely different concepts.

The attitude of EAU Guidelines towards antibiotic treatment of urogenital infections in the context of infertility is rather reserved or sceptical. They assume (i) MAGIs are not clearly associated with impaired natural conception (level of evidence: 3), (ii) the antibiotic treatment leads only to pathogen eradication, which, however, does not lead to the normalization of the inflammatory environment and cannot reverse functional deficits and anatomical abnormalities (level of evidence: 2a), (iii) the antibiotic treatment may improve sperm quality, which necessarily does not lead to increased pregnancy rate (level of evidence: 2a), and finally (iv) current data do not allow to draw any recommendation regarding impact of antibiotic and antioxidant treatment on infertility in males with leukocytospermia (level of evidence: 3) [1].

For those reasons, EAU Guidelines suggest only two recommendations, namely:

• MAGIs should be treated, as it may improve quality of sperm; however, it does not necessarily lead to increased pregnancy rate (strength rating: weak)

• Patients with epididymitis should be advised to test their sexual partners for sexually transmitted disease, since the most likely causal pathogens are *Neisseria gonorrhoeae* or *Chlamydia trachomatis* (strength rating: strong).

The second approach assigns a detrimental impact of urogenital infections on male fertility, which renders an adequate antibiotic or anti-inflammatory treatment mandatory. This proactive attitude is held, for example, by joint guidelines of three national gynaecological and obstetric societies, namely by the German (Deutsche Gesellschaft für Gynäkologie und Geburtshilfe, DGGG), Austrian (Österreichische Gesellschaft für Gynäkologie und Geburtshilfe, OEGGG), and Swiss (Schweizerische Gesellschaft für Gynäkologie und Geburtshilfe, SGGG) societies [5].

The primary aim of this study was to assess the impact of antibiotic treatment of asymptomatic urogenital infections caused by *Ureaplasma urealyticum*, *Mycoplasma hominis*, *Mycoplasma genitalium*, and *Chlamydia trachomatis* on both conventional sperm parameters and sperm apoptosis parameters. The secondary aim was to assess pregnancy outcomes following completion of this treatment.

## 2. Materials and Methods

### 2.1. Study population

Between January 2019 and December 2024, a total of 906 men underwent repeated (at least two consecutive) semen examinations – conventional semen analysis together with flow cytometry sperm apoptosis test (FACS-APO) – at our institution, yielding a total of 2,063 samples. Of these, 679 men and their 613 female partners underwent microbiological testing of first-void morning urine for the presence of *Ureaplasma urealyticum*, *Mycoplasma hominis*, *Mycoplasma genitalium*, and *Chlamydia trachomatis*. No infection was detected in 360 men and their 315 partners. Conversely, infection with at least one of the aforementioned pathogens was detected in 319 couples, of whom 287 underwent simultaneous antibiotic treatment of both partners. A couple was classified as infection-positive if the pathogen was detected in at least one partner, irrespective of whether the other partner was tested, and antibiotic treatment was subsequently initiated in both partners.

We retrospectively identified 265 of these couples (males: age 22.2–62.2 years, mean 36.8 years, standard deviation 6.36 years; females: age 20.7–44.1 years, mean 33.4 years, standard deviation 4.93 years), examined at our institution for infertility lasting 12 months or longer, or a shorter duration in women older than 35 years, in whom a follow-up conventional semen analysis with FACS-APO was performed 0–9 months after completion of antibiotic treatment. Infection was classified as asymptomatic based on the absence of dysuria, urinary urgency, and urethral or vaginal discharge on clinical assessment. Couples were excluded if no urogenital infection was confirmed by first-void morning urine microbiological testing – whether because testing was not performed or because the result was negative – or if a follow-up semen analysis was not obtained within the predefined 0–9-month interval.

### 2.2. Baseline assessment

Baseline assessment included clinical examination of the female partner by a reproductive gynaecologist, together with conventional semen analysis and FACS-APO in the male partner. Both partners also underwent microbiological testing of first-void urine for the presence of *Ureaplasma urealyticum*, *Mycoplasma hominis*, *Mycoplasma genitalium*, and *Chlamydia trachomatis*.

#### 2.2.1. Conventional sperm analysis and flow cytometry sperm apoptosis test (FACS-APO)

A conventional semen analysis of the male partner was performed after 2–7 days of sexual abstinence, according to the WHO laboratory manual for the examination and processing of human semen (5^th^ edition until July 2021, 6^th^ edition from August 2021 onward). Physical characteristics (macroscopic appearance, liquefaction, viscosity, odour, and pH) were documented only when abnormal. The semen was weighed to establish semen volume. A 10 μL aliquot semen was loaded on a sperm counting chamber to assess sperm concentration, total sperm count, and progressive motility for all samples. Total motility was documented from August 2021 onward. Sperm morphology was not consistently assessed throughout the 6-year study period and was therefore not included in the analysis.

In men with abnormal conventional semen parameters, a subsequent clinical examination by an andrologist was performed.

FACS-APO was performed using the ApoFlowEx Kit (Exbio, Prague, Czech Republic) to detect phosphatidylserine externalization via Annexin V-fluorescein isothiocyanate (FITC), and propidium iodide (PI) staining, classifying spermatozoa into three gates: viable, early apoptotic, and late apoptotic/necrotic. A detailed description of the method has been published previously by our group [6].

Based on our previous findings that FACS-APO can identify men unlikely to have elevated sperm DNA fragmentation – thanks to its negative predictive value (NPV) reaching 83.8% [6] – we also compared pre- and post-treatment calculated sperm DNA fragmentation, derived from the previously published formula: sperm DNA fragmentation [%] = 23 + 0.32 × age [years] − 0.35 × viable sperm [%].

The semen examinations were performed in the Laboratory for Embryology and the Laboratory for Immunology, respectively, both belonging to IVF Clinic GENNET, Prague, Czech Republic.

#### 2.2.2. Microbiological assessment

Microbiological testing of first-void morning urine included culture-based detection of *Ureaplasma urealyticum* and *Mycoplasma hominis*, and PCR-based molecular detection of *Mycoplasma genitalium* and *Chlamydia trachomatis*. Antimicrobial susceptibility testing was performed for culture-positive organisms (*Ureaplasma urealyticum* and *Mycoplasma hominis*); susceptibility data were not available for organisms detected by PCR (*Mycoplasma genitalium* and *Chlamydia trachomatis*).

#### 2.2.3. Partner infection concordance

To assess concordance of urogenital infection status between male and female partners, this analysis was restricted to the 244 couples (of the total 265) in whom both partners underwent complete microbiological testing for all four investigated pathogens (*Ureaplasma urealyticum*, *Mycoplasma hominis*, *Mycoplasma genitalium*, and *Chlamydia trachomatis*), as concordance could not be reliably determined when testing was incomplete for either partner. A partner was classified as infection-positive if at least one of the four pathogens was detected, and as infection-negative only if testing for all four pathogens was completed and yielded negative results. Conditional probabilities of partner positivity (i.e., the probability of female positivity given male positivity, and vice versa) were calculated as simple proportions among couples with complete bilateral testing.

### 2.3. Treatment

Simultaneous antibiotic treatment was initiated for both partners in all couples immediately upon detection of the aforementioned bacteria. The standard regimen consisted of doxycycline 100 mg orally every 12 hours for 10 days (241 cases). Alternative antibiotics – azithromycin (12 cases), levofloxacin (10 cases), moxifloxacin (1 case), or clarithromycin (1 case) – were used only in cases of known allergy, newly developed intolerance, or documented resistance. Couples were instructed to avoid concomitant intake of doxycycline with dairy products or dietary supplements containing Mg²⁺ or Zn²⁺ in order to ensure treatment efficacy.

### 2.4. Follow-up

A follow-up conventional semen analysis with FACS-APO was performed in men 0–9 months after completion of antibiotic treatment.

In addition to the effect of antibiotic treatment on semen and FACS-APO parameters, we assessed whether pregnancy was achieved within 12 months of completing antibiotic treatment, and by what means – spontaneously, following intrauterine insemination (IUI), following in vitro fertilization (IVF) using the couple’s own oocytes, or following IVF with donor oocytes. For descriptive purposes, pregnancies were further categorized according to whether they occurred within 0–6 months or 7–12 months after completion of antibiotic treatment. Pregnancy was confirmed by ultrasound, performed either by a gynaecologist at our institution or by the patient’s registering gynaecologist, with this information subsequently reported by the patient. Spontaneous miscarriage occurring after spontaneous conception or after IVF within this 12-month period was likewise documented.

Couples without a documented pregnancy were further classified as: no pregnancy despite IVF treatment; no pregnancy without IVF having been pursued; or lost to follow-up (i.e., pregnancy status could not be ascertained). The pregnancy rate was calculated both as an intention-to-treat rate (using the total cohort, including couples lost to follow-up, as the denominator) and as a per-protocol rate (excluding couples lost to follow-up from the denominator).

### 2.5. Statistical Analysis

Continuous variables are presented as mean ± standard deviation (SD) and median (interquartile range, IQR), as appropriate. Paired baseline and follow-up measurements were available for 265 couples for each of the assessed semen and FACS-APO parameters, with the exception of total motility, which was available for only 130 cases.

For each parameter, the normality of the paired differences between baseline and follow-up values was assessed using the Shapiro-Wilk test. Where the assumption of normality was met, a paired-samples t-test was used to compare baseline and follow-up values, with effect size expressed as Cohen’s d and further supported by a Bayes factor (BF₁₀) and post hoc statistical power. Where the assumption of normality was violated, the non-parametric Wilcoxon signed-rank test was used instead, with effect size expressed as the rank-biserial correlation (RBC) and the common language effect size (CLES). A two-sided p-value < 0.05 was considered statistically significant for all paired comparisons.

To evaluate whether the magnitude of change in semen and FACS-APO parameters differed according to the timing of the follow-up examination, couples were stratified into three subgroups based on the interval between completion of antibiotic treatment and follow-up semen analysis (0–3, 4–6, and 7–9 months, respectively). For each parameter, normality within each subgroup was assessed using the Shapiro-Wilk test, and homogeneity of variances across subgroups was assessed using Levene’s test. Where both assumptions were met, a one-way analysis of variance (ANOVA) was used to compare the three subgroups; otherwise, the non-parametric Kruskal-Wallis test was used. Where the Kruskal-Wallis test indicated a statistically significant difference, pairwise post hoc comparisons were performed using Dunn’s test with Holm correction for multiple comparisons.

Statistical analyses were performed in Python (pandas, scipy, and pingouin libraries). Statistical significance was set at p < 0.05 throughout.

## 3. Results

### 3.1. Semen and FACS-APO parameters before and after antibiotic treatment

Paired baseline and follow-up measurements were available for 265 couples for most reported parameters. Total sperm motility was recorded systematically only from July 2020 onward and was therefore available for a smaller subset of the cohort (130 of 265 pairs).

The results of the paired comparisons are summarized in Table 1 and in Figure 2.

**Table 1.** Semen and FACS-APO parameters at baseline and follow-up (paired comparison)

| Parameter | n | Baseline, mean ± SD<br>(median) | Follow-up, mean<br>± SD (median) | Test | Statistic | p-value | Effect size |
| --- | --- | --- | --- | --- | --- | --- | --- |
| **Semen volume (mL)** | 265 | 3.35 ± 1.69 (3.0) | 3.51 ± 1.95 (3.1) | Wilcoxon | W = 12,149.0 | 0.123 | RBC = -0.116 |
| **Sperm concentration**<br>**($\times 10^6$/mL)** | 265 | 24.62 ± 28.85 (13.0) | 27.70 ± 27.47<br>(18.0) | Wilcoxon | W = 13,006.5 | 0.00371 | RBC = -0.209 |
| **Total sperm count**<br>**($\times 10^6$)** | 265 | 78.80 ± 103.02 (36.4) | 90.23 ± 102.86<br>(60.0) | Wilcoxon | W = 13,553.5 | 0.00112 | RBC = -0.231 |
| **Total motility (%)** | 130 | 39.83 ± 16.96 (40.0) | 43.80 ± 15.73<br>(42.9) | Paired t-test | t = -3.09 | 0.00243 | d = 0.243 |
| **Progressive motility**<br>**(%)** | 265 | 31.06 ± 18.10 (29.0) | 35.44 ± 18.63<br>(33.3) | Paired t-test | t = -4.11 | 0.000053 | d = 0.238 |
| **Viable sperm (%)** | 265 | 39.13 ± 11.99 (40.0) | 42.36 ± 12.41<br>(42.5) | Wilcoxon | W = 11,733 | 0.000002 | RBC = -0.334 |
| **Early apoptotic**<br>**sperm (%)** | 265 | 16.39 ± 8.60 (14.8) | 16.08 ± 8.29<br>(14.1) | Wilcoxon | W = 17,335.5 | 0.901 | RBC = 0.009 |
| **Late apoptotic/necrotic sperm (%)** | 265 | 44.50 ± 12.26 (44.5) | 41.57 ± 11.54 (41.0) | Wilcoxon | W = 12,541.0 | 0.000047 | RBC = 0.288 |
| **Total apoptotic sperm (%)** | 265 | 60.88 ± 11.99 (60.0) | 57.64 ± 12.40 (57.5) | Wilcoxon | W = 11,731.5 | 0.000002 | RBC = 0.334 |
| **Calculated sperm DNA fragmentation** | 265 | 21.04 ± 4.79 (21.0) | 19.91 ± 4.84 (19.9) | Wilcoxon | W = 11,468.0 | 0.000002<br>7 | RBC = 0.349 |

**Figure 1.**
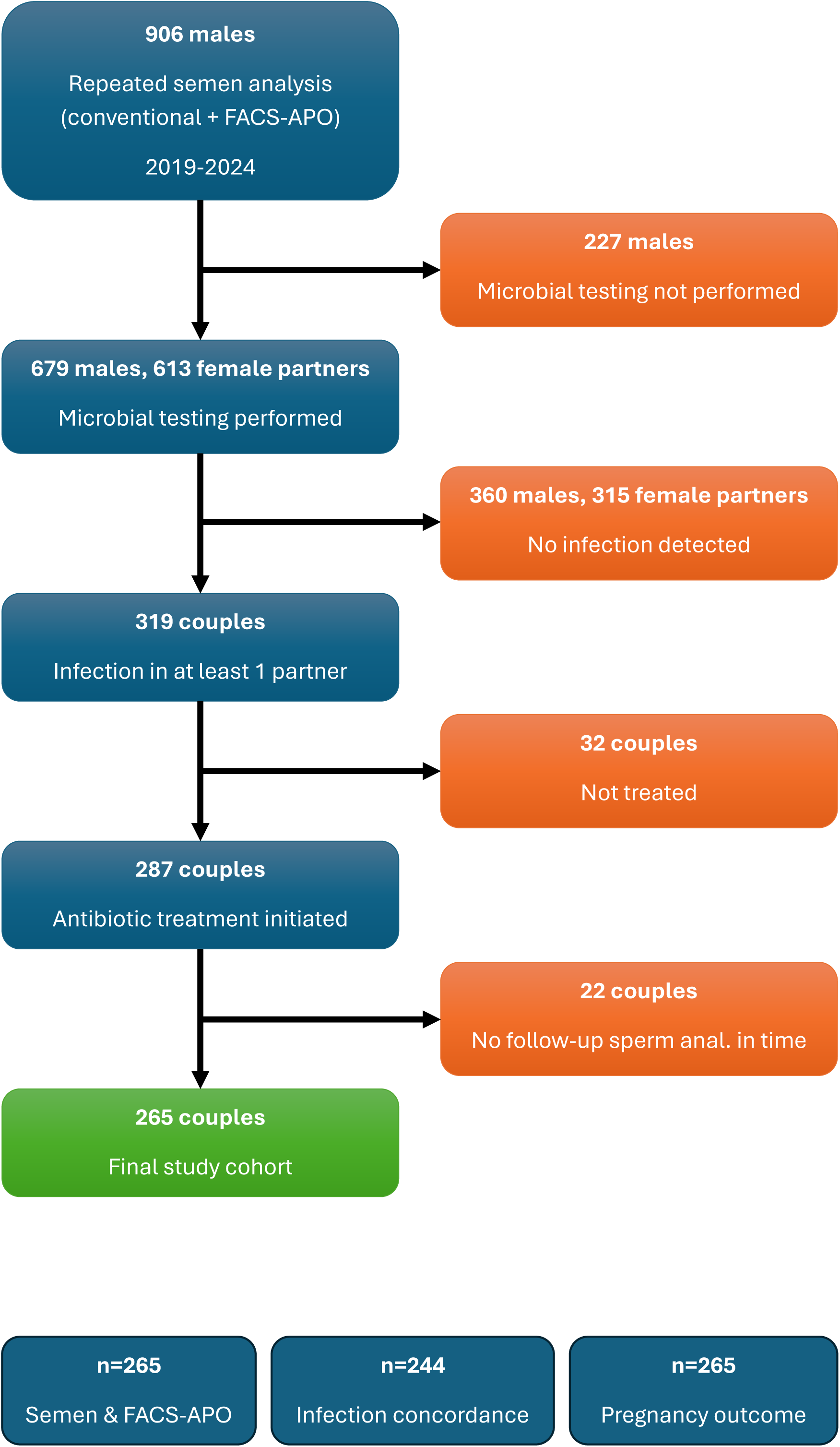
Flow diagram showing selection of the study cohort from initial semen examination through microbiological testing, antibiotic treatment, and the subsets used for follow-up semen analysis, partner infection concordance, and pregnancy outcome analyses.

**Figure 2.**
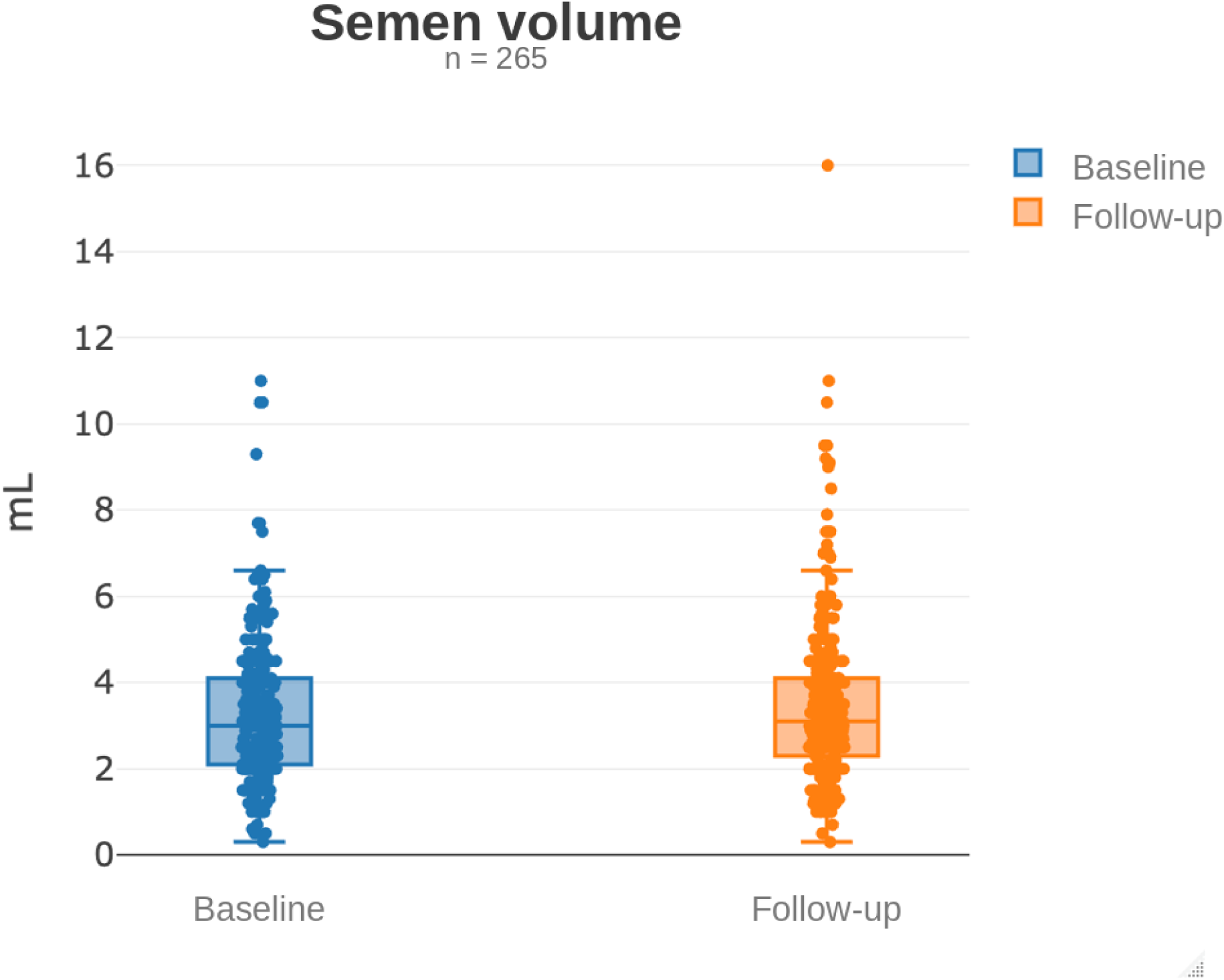
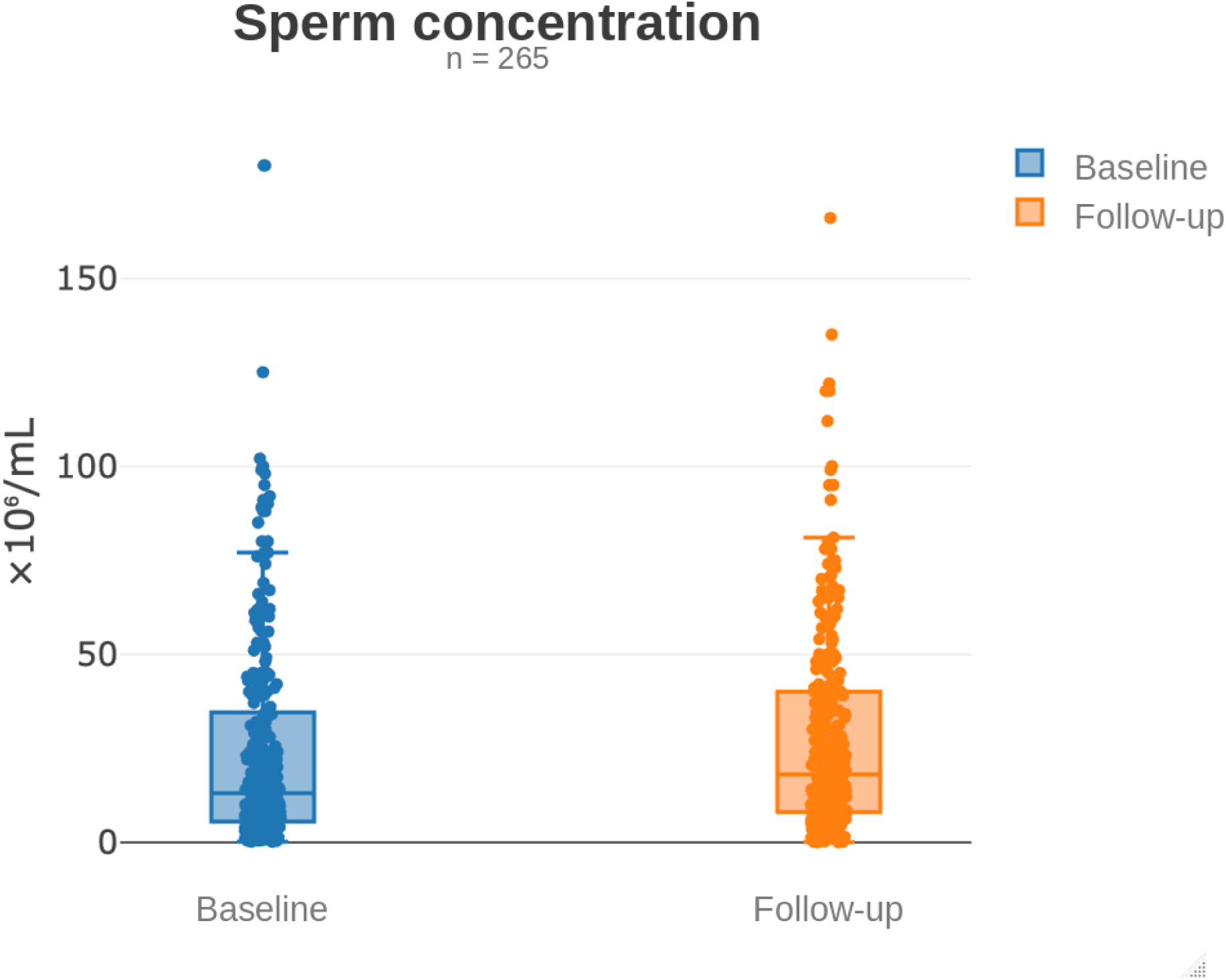
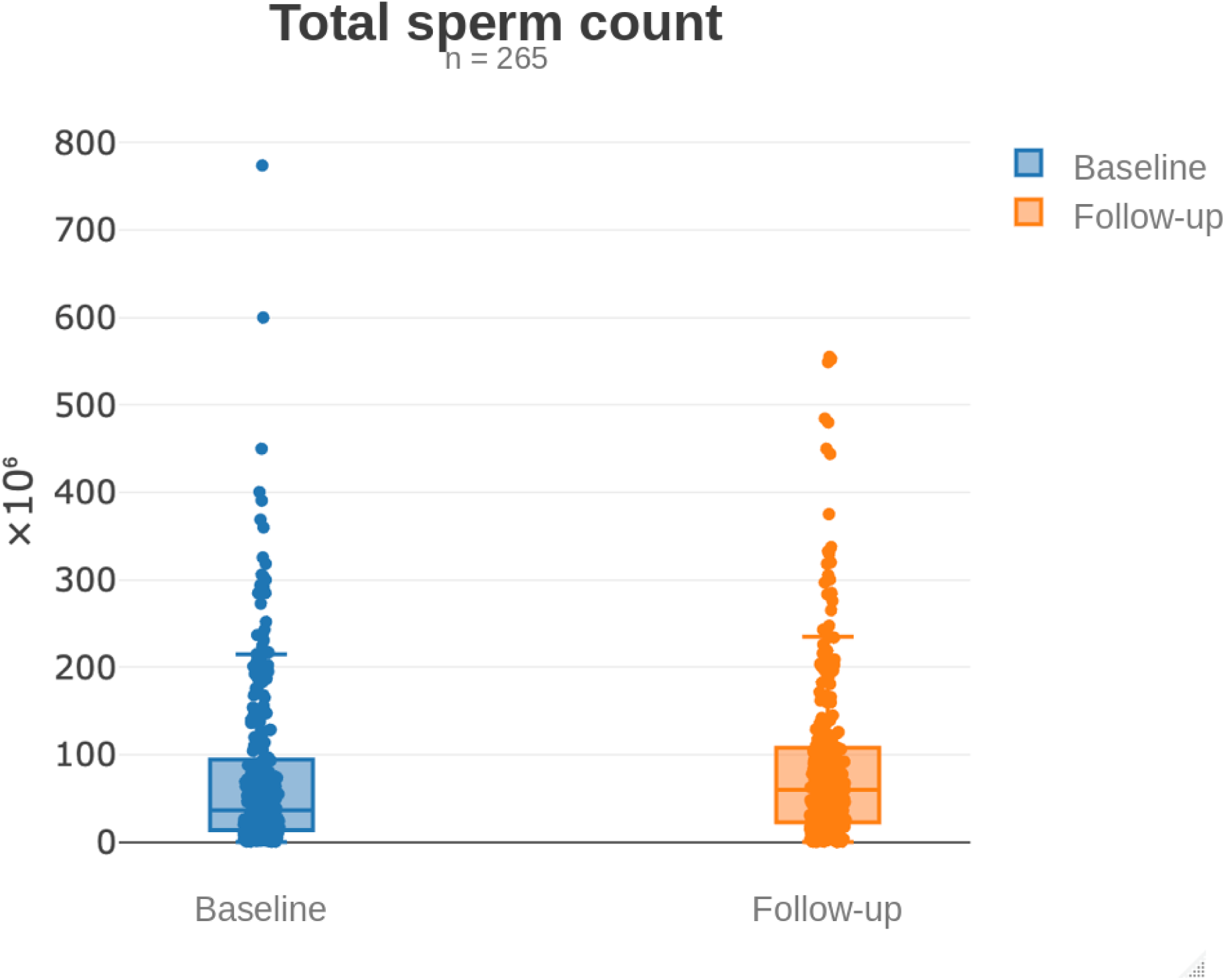
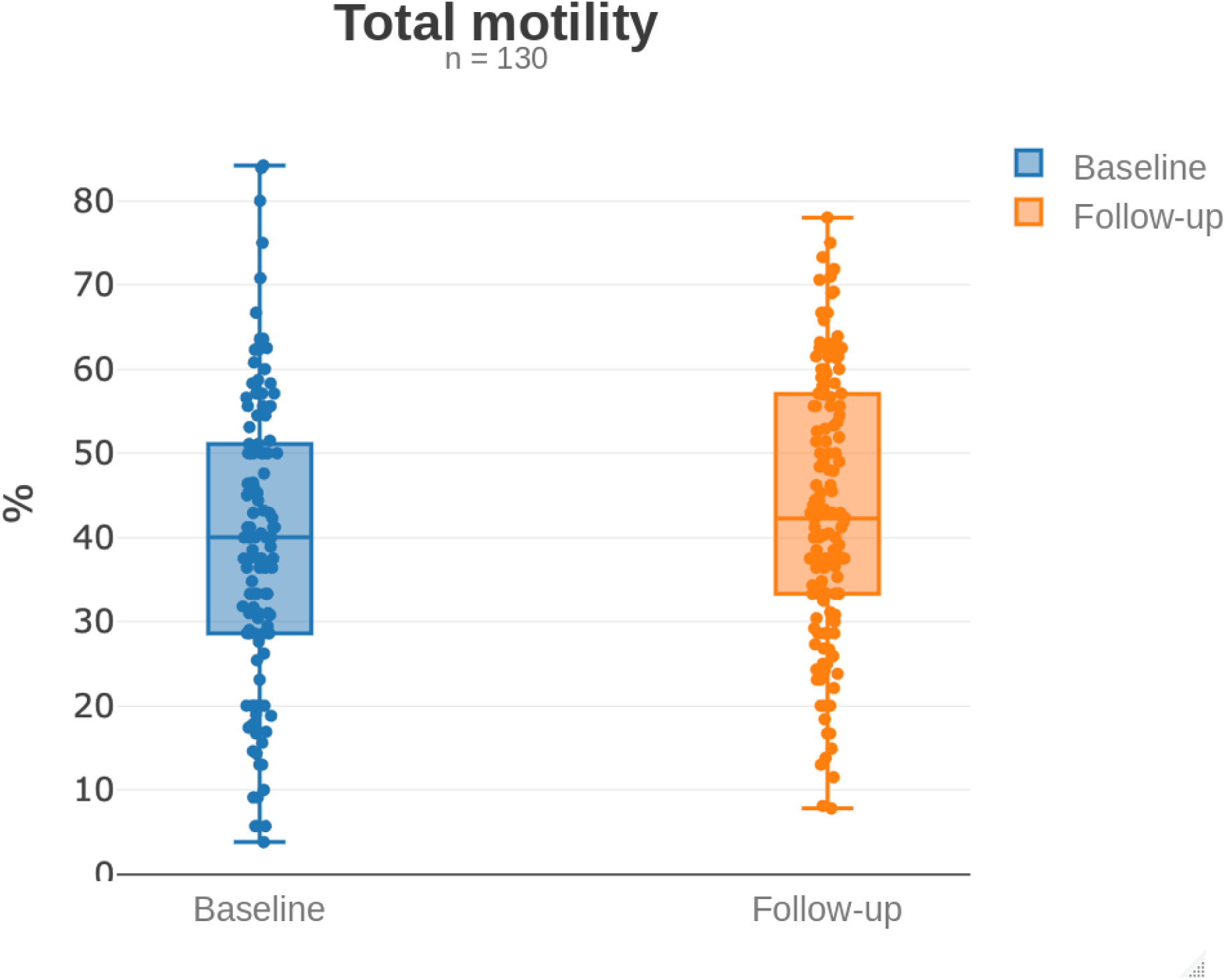
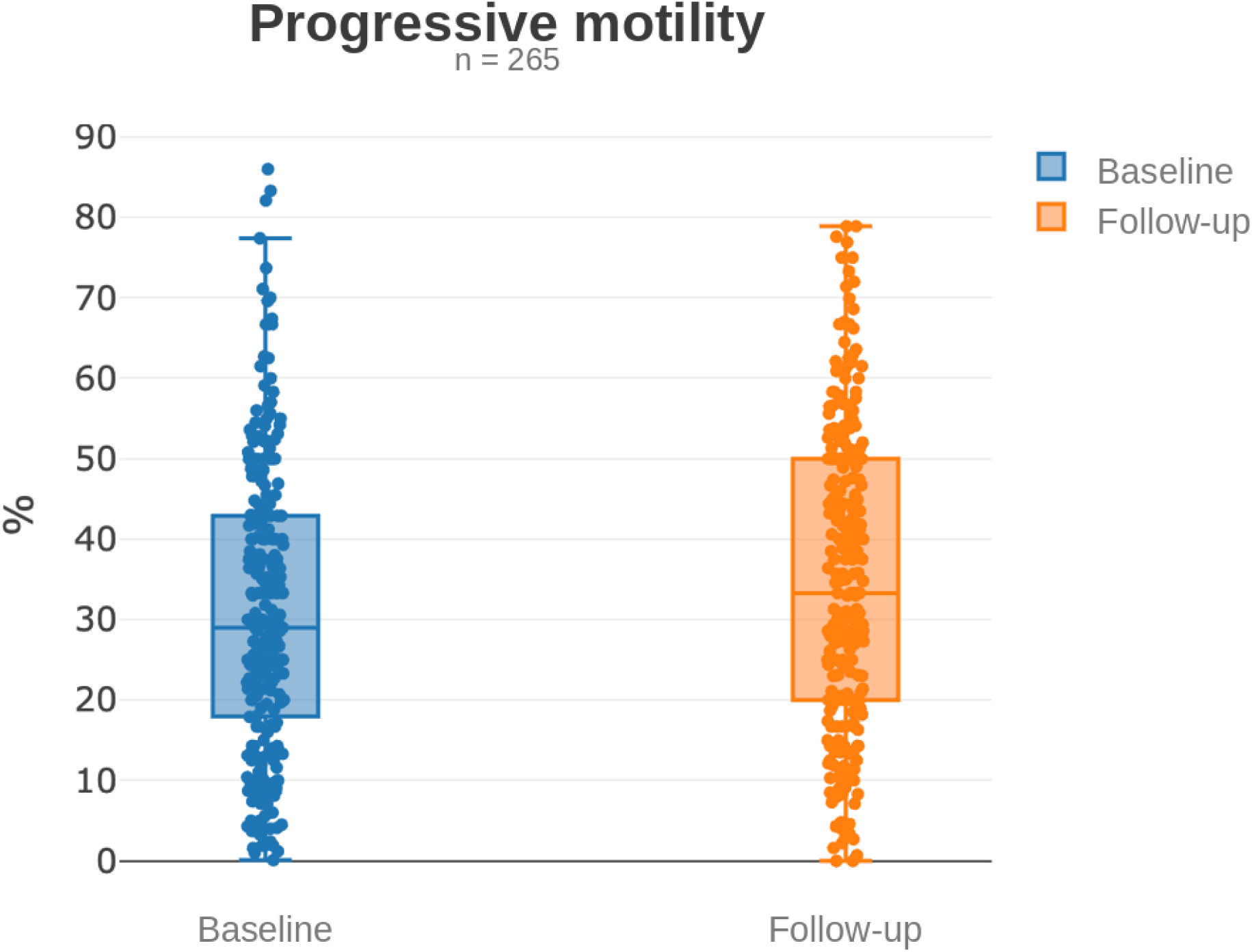
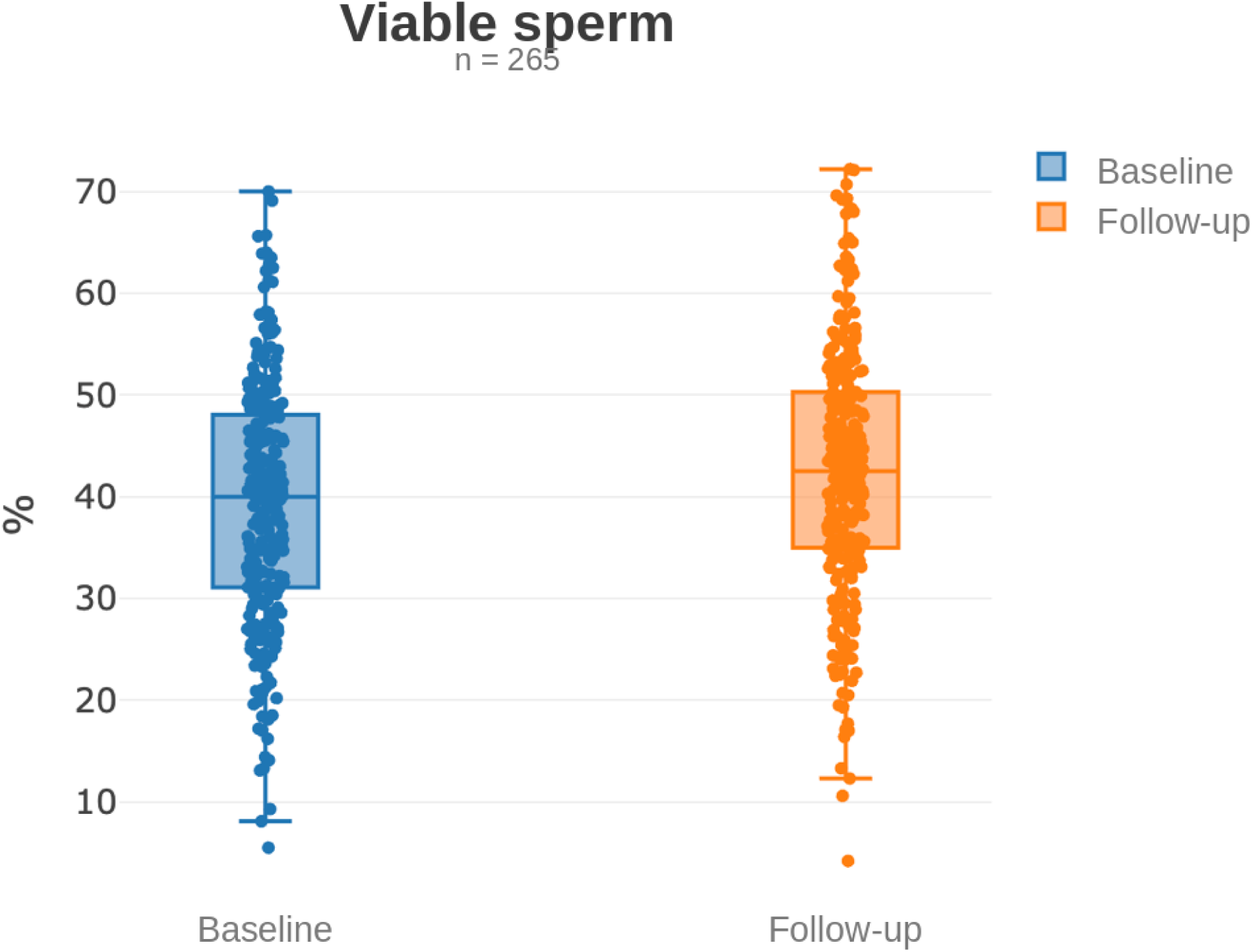
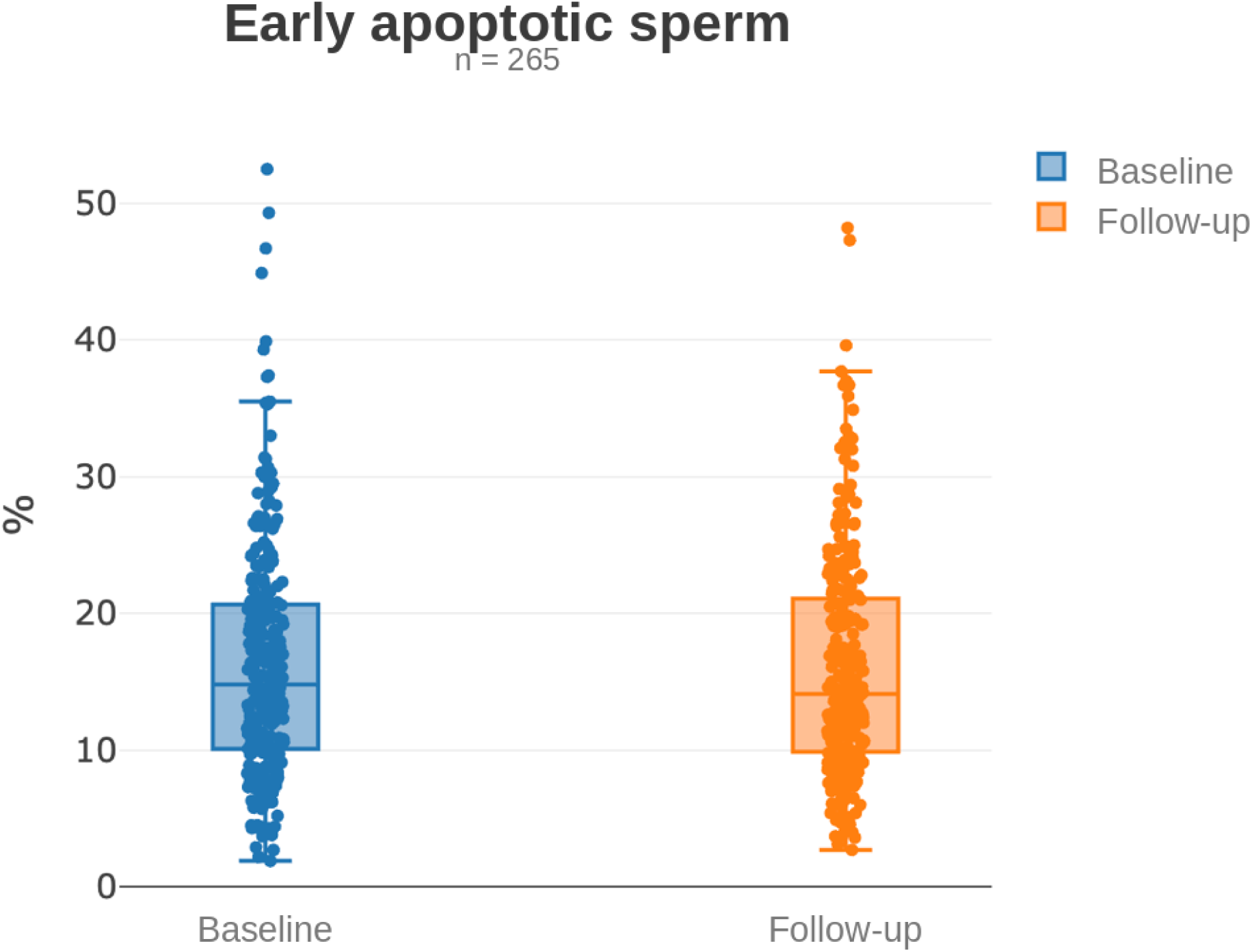
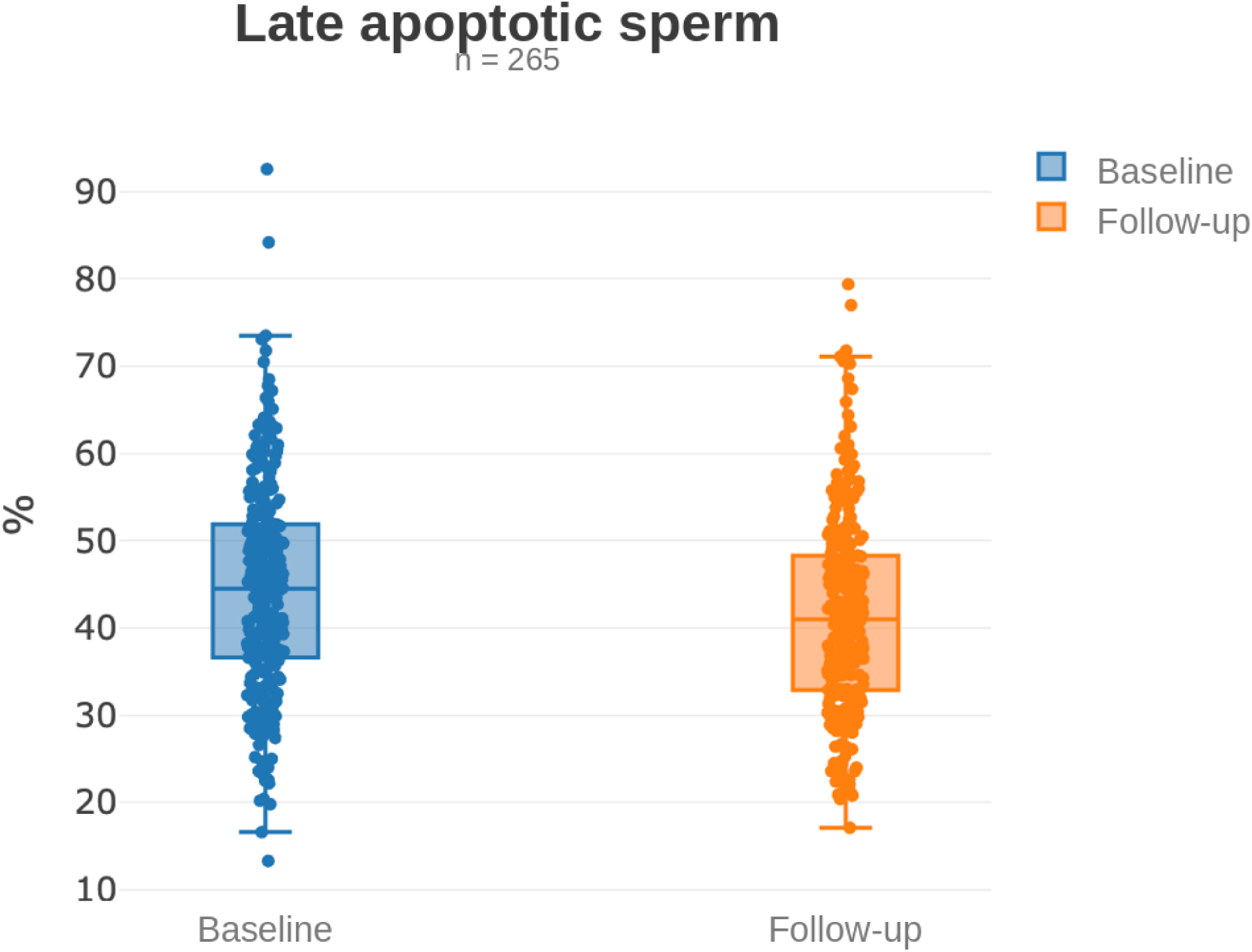
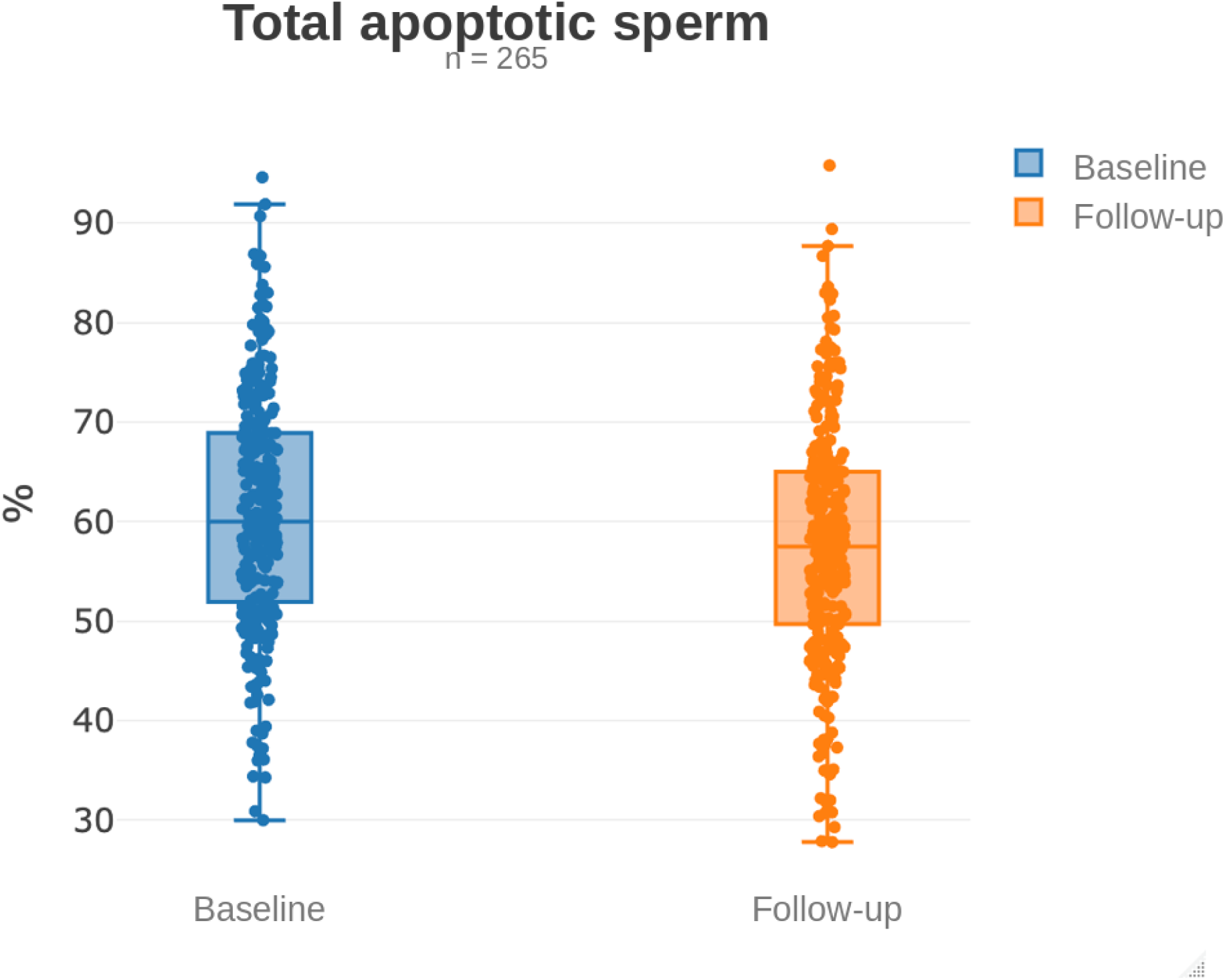
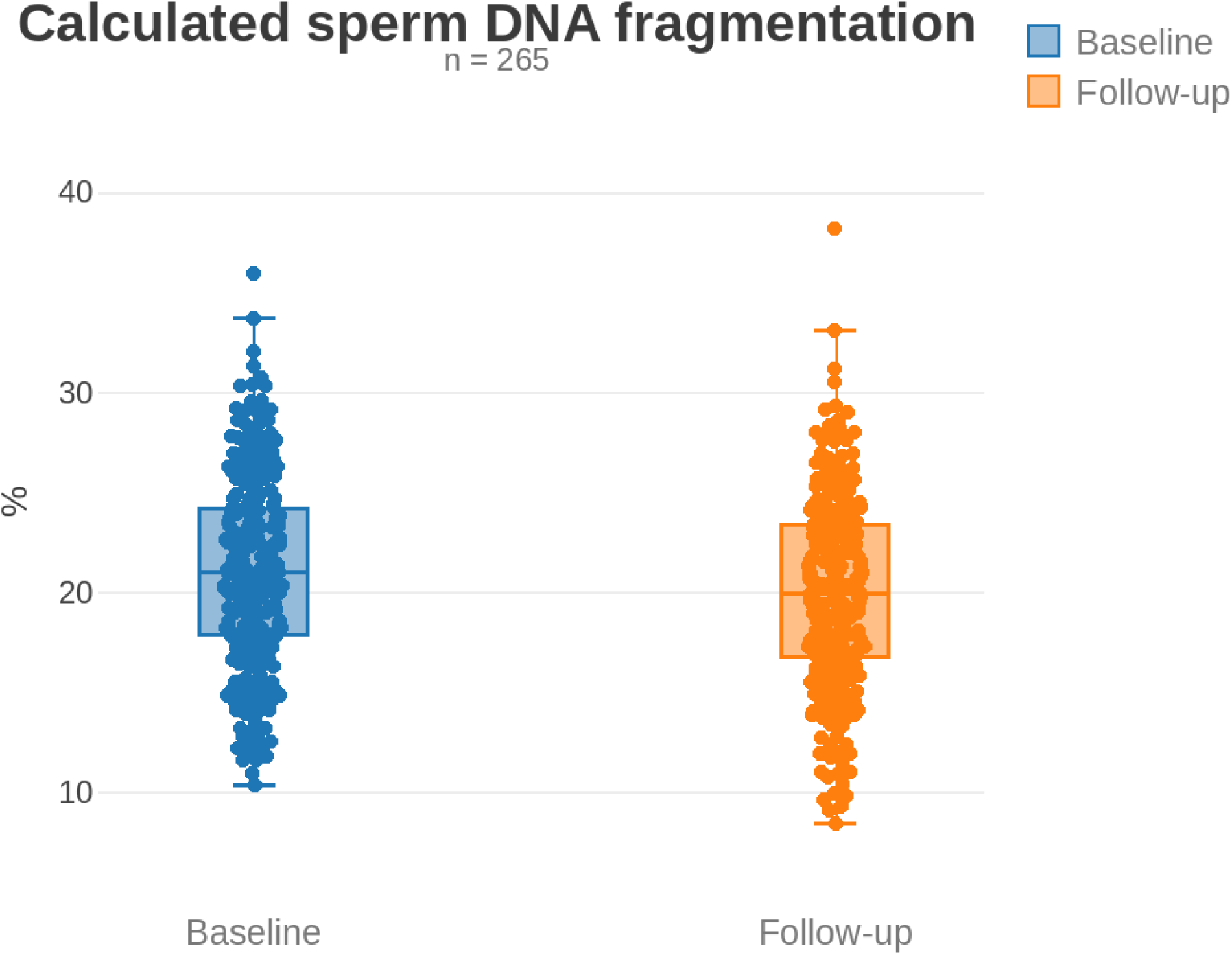
Semen and FACS-APO parameters before and after antibiotic treatment. Box plots show (A) semen volume, (B) sperm concentration, (C) total sperm count, (D) total motility, (E) progressive motility, (F) viable sperm, (G) early apoptotic sperm, (H) late apoptotic / necrotic sperm, (I) total apoptotic sperm, and (J) calculated sperm DNA fragmentation, at baseline and at follow-up after completion of antibiotic treatment. Boxes indicate the interquartile range (IQR), with the horizontal line representing the median; whiskers extend to the most extreme value within 1.5 × IQR of the box. Individual data points are shown as dots. n = 265 for all parameters except total motility (n = 130; recorded systematically only from July 2020 onward – see Methods).

#### Semen volume

Semen volume did not change significantly between baseline and follow-up (mean 3.35 mL vs. 3.51 mL; Wilcoxon signed-rank test, W = 12,149.0, p = 0.123, RBC = −0.116), supporting the assumption that the subsequently observed improvements in sperm count and concentration were not attributable to differences in ejaculate volume between the two collections.

#### Sperm concentration

Sperm concentration increased significantly between baseline and follow-up (mean 24.6 × 10⁶/mL vs. 27.7 × 10⁶/mL; median 13.0 vs. 18.0 × 10⁶/mL; W = 13,006.5, p = 0.00371, RBC = −0.209).

#### Sperm count

The total sperm count increased significantly (mean 78.8 × 10⁶ vs. 90.2 × 10⁶; median 36.4 × 10⁶ vs. 60.0 × 10⁶; W = 13,553.5, p = 0.00112, RBC = −0.231).

#### Sperm motility

Total motility increased significantly between baseline and follow-up (130 pairs; mean 39.8% vs. 43.8%; paired t-test, t = −3.09, df = 129, p = 0.00243, 95% CI of the difference −6.51 to −1.43 percentage points, Cohen’s d = 0.243, BF₁₀ = 8.96, power = 78%). Progressive motility likewise increased significantly (265 pairs; mean 31.1% vs. 35.4%; t = −4.11, df = 263, p = 0.000053, 95% CI −6.48 to −2.28 percentage points, Cohen’s d = 0.238, BF₁₀ = 226.85, power = 97%).

#### Apoptosis-related parameters (FACS-APO)

The proportion of viable spermatozoa increased significantly between baseline and follow-up (mean 39.1% vs. 42.4%; median 40.0% vs. 42.5%; W = 11,733, p = 0.000002, RBC = −0.334), and the proportion of late apoptotic / necrotic spermatozoa decreased significantly (mean 44.5% vs. 41.6%; W = 12,541.0, p = 0.000047, RBC = 0.288), together resulting in a significant decrease in the total proportion of apoptotic spermatozoa (mean 60.9% vs. 57.6%; W = 11,731.5, p = 0.000002, RBC = 0.334). The proportion of early apoptotic spermatozoa did not change significantly (mean 16.4% vs. 16.1%; W = 17,335.5, p = 0.901, RBC = 0.009).

#### Calculated sperm DNA fragmentation

Calculated sperm DNA fragmentation decreased significantly between baseline and follow-up (mean 21.0% vs. 19.9%; W = 11,468.0, p = 0.0000027, RBC = 0.349), consistent with the concurrent decrease in total apoptotic sperm.

### 3.2. Partner infection concordance

Of the 265 couples included in the study, both partners underwent complete microbiological testing for all four investigated pathogens (*Ureaplasma urealyticum*, *Mycoplasma hominis*, *Mycoplasma genitalium*, and *Chlamydia trachomatis*) in 244 couples (92.1%); the remaining 21 couples (7.9%) were excluded from the concordance analysis because testing was incomplete for at least one partner. Among these 21 couples, the male partner had already tested positive in 15 (71%), while microbiological testing of the female partner was incomplete or, in 8 cases, not performed at all – reflecting a clinical decision to omit full testing of the female partner once infection had been confirmed in the male partner. This non-random pattern of missing data may have led to a slight underestimation of true partner concordance in the present analysis.

Among the 244 fully tested couples, at least one of the four pathogens was detected in 153 men (62.7%) and 218 women (89.3%). *Ureaplasma urealyticum* was by far the most frequently detected pathogen in both men (141/244, 57.8%) and women (214/244, 87.7%), and both partners tested positive for *Ureaplasma urealyticum* concurrently in 123 couples (50.4%). *Mycoplasma hominis* was detected in 13 men (5.3%) and 26 women (10.7%), with concurrent positivity in 5 couples (2.0%); *Chlamydia trachomatis* was detected in 9 men (3.7%) and 3 women (1.2%), with concurrent positivity in 2 couples (0.8%); and *Mycoplasma genitalium* was detected in 2 men (0.8%) and 1 woman (0.4%), with no couple positive in both partners concurrently (Table 2). Co-infection with two or more pathogens was observed in 12 men (4.9%) and 26 women (10.7%).

**Table 2.** Pathogen positivity and partner concordance among the 244 fully tested couples.

| **Pathogen** | **Male positive, n (%)** | **Female positive, n (%)** | **Both partners positive, n (%)** |
| --- | --- | --- | --- |
| *Ureaplasma urealyticum* | 141 (57.8%) | 214 (87.7%) | 123 (50.4%) |
| *Mycoplasma hominis* | 13 (5.3%) | 26 (10.7%) | 5 (2.0%) |
| *Chlamydia trachomatis* | 9 (3.7%) | 3 (1.2%) | 2 (0.8%) |
| *Mycoplasma genitalium* | 2 (0.8%) | 1 (0.4%) | 0 (0.0%) |
| **Any pathogen** | 153 (62.7%) | 218 (89.3%) | 127 (52.0%) |

At the couple level, both partners tested positive for at least one pathogen in 127 couples (52.0%), only the male partner was positive in 26 couples (10.7%), and only the female partner was positive in 91 couples (37.3%); no couple tested negative in both partners, consistent with the study’s inclusion criteria. Among couples in which the male partner tested positive, the female partner was also positive in 127 of 153 couples (83.0%). Conversely, among couples in which the female partner tested positive, the male partner was also positive in 127 of 218 couples (58.3%).

### 3.3. Effect of follow-up timing on the magnitude of change

When couples were stratified by the interval between completion of antibiotic treatment and follow-up semen analysis (0–3, 4–6, and 7–9 months), the magnitude of change (Δ) differed significantly across subgroups for sperm concentration (Kruskal-Wallis H = 11.65, p = 0.0030) and total sperm count (H = 10.07, p = 0.0065). For both parameters, post hoc pairwise comparisons (Dunn’s test, Holm-corrected) showed significant differences between the 0–3-month and 7–9-month subgroups (all p ≤ 0.008, Hedges’ g ≈ 1.05) and between the 4–6-month and 7–9-month subgroups (all p ≤ 0.028, Hedges’ g ≈ 0.79–1.05), while the 0–3-month and 4–6-month subgroups did not differ significantly. Descriptively, the mean increase in total sperm count was largest in the 0–3-month subgroup (+16.5 × 10⁶, n = 200) and smaller in the 4–6-month subgroup (+10.3 × 10⁶, n = 54), whereas the 7–9-month subgroup (n = 11) showed a mean decrease (−75.1 × 10⁶); a similar pattern was observed for sperm concentration, with a mean increase of +4.6 × 10⁶/mL (0–3 months), +1.6 × 10⁶/mL (4–6 months), and a mean decrease of −17.3 × 10⁶/mL (7–9 months). No significant effect of follow-up timing was observed for the remaining parameters (all p > 0.05).

### 3.4. Pregnancy outcomes

Among the 265 couples, pregnancy was achieved in 127 (47.9%; per-protocol rate excluding 27 couples lost to follow-up: 127/238, 53.4%) within 12 months of completing antibiotic treatment (Table 3). Of these, 21 pregnancies (16.5%) occurred spontaneously, 14 (11.0%) following IUI, 72 (56.7%) following IVF using the couple’s own oocytes, and 2 (1.6%) following IVF with donor oocytes; a further 9 pregnancies following spontaneous conception and 9 following IVF ended in miscarriage. The majority of pregnancies (91/127, 71.7%) occurred within the first 6 months after treatment completion. Among the 138 couples without a documented pregnancy, 66 (47.8%) had undergone IVF without success, 45 (32.6%) had not conceived spontaneously within 12 months and had not pursued IVF, and 27 (19.6%) were lost to follow-up.

**Table 3.**
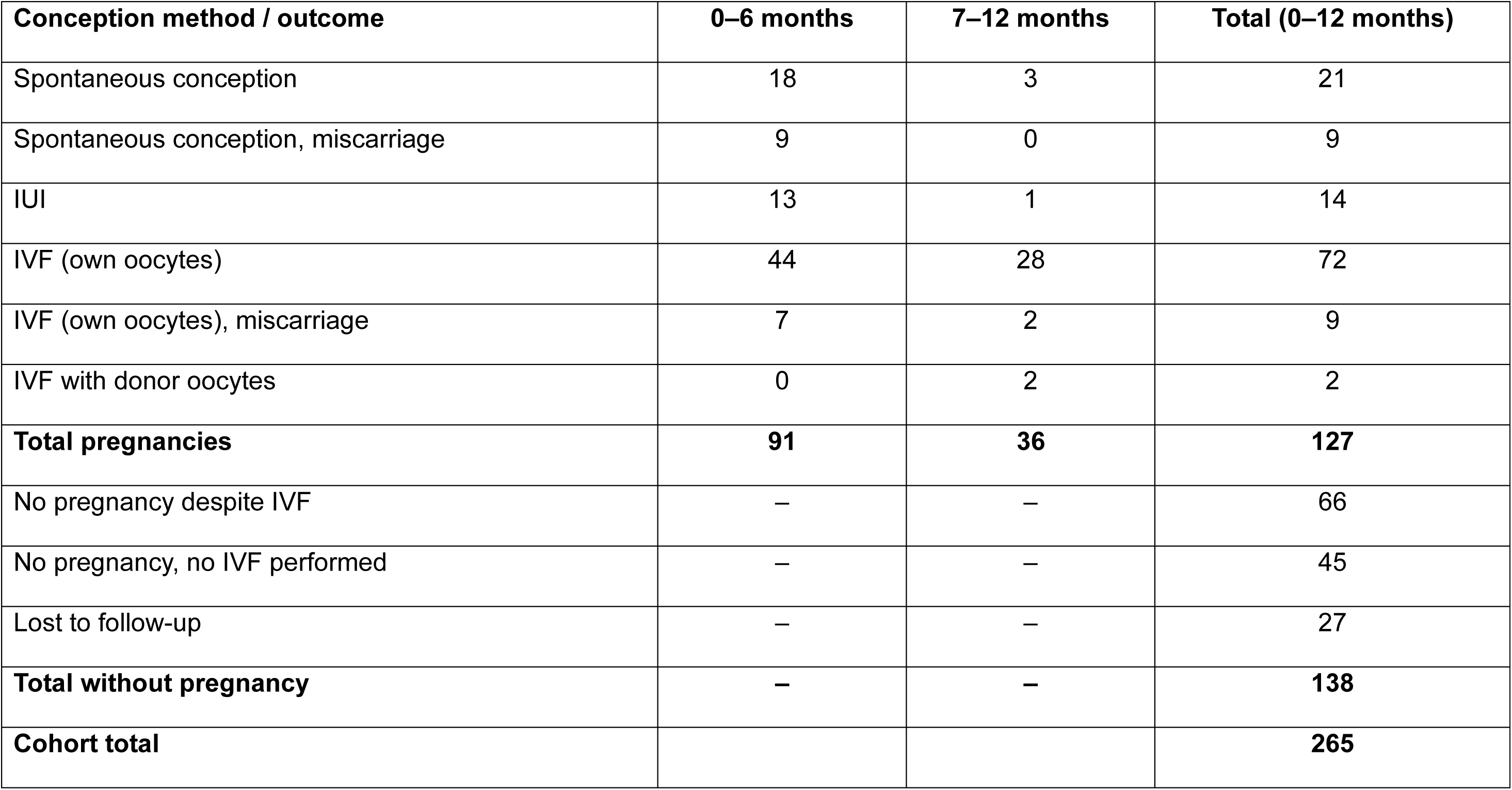
Pregnancy outcomes following antibiotic treatment, by conception method and time since completion of treatment.

| Conception method / outcome | 0–6 months | 7–12 months | Total (0–12 months) |
| --- | --- | --- | --- |
| Spontaneous conception | 18 | 3 | 21 |
| Spontaneous conception, miscarriage | 9 | 0 | 9 |
| IUI | 13 | 1 | 14 |
| IVF (own oocytes) | 44 | 28 | 72 |
| IVF (own oocytes), miscarriage | 7 | 2 | 9 |
| IVF with donor oocytes | 0 | 2 | 2 |
| **Total pregnancies** | **91** | **36** | **127** |
| No pregnancy despite IVF | – | – | 66 |
| No pregnancy, no IVF performed | – | – | 45 |
| Lost to follow-up | – | – | 27 |
| **Total without pregnancy** | **–** | **–** | **138** |
| **Cohort total** |  |  | **265** |

## 4. Discussion

Although the impact of asymptomatic urogenital infections on male and female fertility has been widely studied, the level of evidence is generally low, and thus remaining a controversial issue [3,7–9]. Given the physiology of human reproduction, as discussed in the Limitations section, truly high-quality data on this topic can hardly be expected.

The negative impact of *Chlamydia trachomatis* [10,11] or *Neisseria gonorrhoeae* [12–14] on both male and female fertility has been advocated by many authors. A systematic review and meta-analysis, focusing specifically on genital mycoplasmas and ureaplasmas (collectively classified within the class *Mollicutes* – from Latin *mollis* = soft, *cutis* = skin, denoting bacteria lacking a cell wall), suggested that this association might be species-specific: a significant relationship with male infertility was found for *Ureaplasma urealyticum* and *Mycoplasma hominis*, whereas no such association was observed for *Ureaplasma parvum* or *Mycoplasma genitalium* [15]. Given the pronounced imbalance in pathogen distribution within the cohort – *Ureaplasma urealyticum* was detected in at least one partner in 247 of 265 couples (93.2%), whereas *Mycoplasma genitalium*, whose causal role in infertility has not been unequivocally confirmed, was detected in only 4 couples (once in combination with *Ureaplasma urealyticum*) – a meaningful statistical comparison between pathogen-specific subgroups was not feasible, and we therefore decided not to evaluate the cohort according to specific bacterial species.

Traditionally, the Meares-Stamey 4-glass test had been used since the 1960s to diagnose chronic prostatitis, enabling precise localization of the source of bacteria. Since the test comprises microbiological cultivation of sequentially collected four body fluids, namely first-void urine (VB1), midstream urine (VB2), expressed prostatic secretions (EPS) after prostate massage, and post-massage urine (VB3) [16], due to its ponderousness, complexity, and last but not least inconvenience for both the patient and the physician, it had often been replaced by simpler and less burdensome 2-glass test requiring only pre-massage (VB2) and post-massage (VB3) urine samples with comparable reproducibility [17].

In contrast to these cultivation-based, multi-sample protocols, modern nucleic acid amplification tests (NAATs) allow reliable pathogen detection from a single specimen. In comparison to urine culture, NAATs show significantly higher sensitivity in diagnosis of asymptomatic urogenital infections in the context of infertile couples (92-99% [18,19] vs. 50-85% [20]), however, without antibiotic susceptibility pattern.

Currently, NAATs are the preferred method for detecting asymptomatic urogenital infections in infertile couples [21,22]. For men, the optimal biological material is first-voided urine, with meatal or urethral swabs being alternatives for *Ureaplasma urealyticum* and *Mycoplasma hominis*. For women, a vaginal swab is preferred, as these bacteria in women typically cause cervicitis; with first-voided urine being an acceptable alternative, however, slightly less sensitive for *Chlamydia trachomatis* and *Mycoplasma genitalium* [20,23,24].

Bacteria causing asymptomatic urogenital infections may colonise non-genital mucoses (i. e., rectal, pharyngeal) [20,25]. Reported rectal colonization rates with *Chlamydia trachomatis* range up to 8–9% in both women and heterosexual men, with pharyngeal colonization reported in approximately 1.6–1.7% of cases [25] – indicating that this phenomenon is not confined to men who have sex with men (MSM), even though most available data on extragenital colonization still originate from this population. A similar pattern has been reported for *Mycoplasma genitalium*: in a cross-sectional STI clinic cohort, extragenital infection was identified in 19.7% of patients after excluding those with concomitant urogenital infection, all of whom were asymptomatic [26]. Data on extragenital colonization with *Ureaplasma urealyticum* remain particularly limited, partly because validated NAAT assays for extragenital specimens are not widely available for this pathogen. Since there are limited data on the prevalence and clinical impact of extragenital colonization specifically in infertile couples, extragenital site testing is generally not recommended in this population unless sexual history indicates risk. Furthermore, in our own cohort, the female partner tested positive for at least one of the investigated pathogens in 83.0% of couples in which the male partner tested positive (Table 2), underscoring the high probability of concurrent infection in the partner even when individual confirmatory testing is impractical or incomplete. Given that apparently pathogen-negative partners may nonetheless harbour an undetected extragenital reservoir of infection, and that our own data demonstrate a high probability of concurrent partner infection, it appears reasonable to treat both partners of a couple with antibiotics as a whole, regardless of individual urogenital test results, in order to prevent persistence or re-transmission of infection [22].

Several studies have described both an improvement in conventional semen parameters and a reduction in reactive oxygen species (ROS) levels following antibiotic treatment of asymptomatic infections caused by *Ureaplasma urealyticum* [27] and *Mycoplasma genitalium* [28]. For *Chlamydia trachomatis*- and *Ureaplasma urealyticum*-related infection specifically, this effect has been shown to be more pronounced 30 days after treatment completion than at 10 days [29], possibly reflecting resolution of acute inflammation rather than a complete turnover of the spermatozoal population, given that a full sperm generation cycle requires substantially longer. In contrast, our study observed a decline in treatment effect in samples collected 6–9 months after antibiotic treatment. However, this subgroup comprised only 11 couples, and reinfection cannot be excluded, as microbiological cure was not confirmed after treatment (see Limitations).

Sperm DNA fragmentation is currently considered the most reliable indicator of male fertilizing potential [30]. Several studies employing heterogeneous designs and methods have linked genitourinary infections caused by *Chlamydia trachomatis*, *Ureaplasma*, and *Mycoplasma* species to increased sperm DNA fragmentation [31,32], while antibiotic treatment has been shown to reduce infection-induced sperm DNA fragmentation [33]. To our knowledge, the present study is the first to systematically monitor apoptosis parameters assessed by a FACS-APO in this context. As we have previously demonstrated, thanks to its high NPV, FACS-APO can be used to select men at low risk of high sperm DNA fragmentation, sparing them the need for complex and generally costly sperm DNA fragmentation testing [6]. Our data have shown that antibiotic treatment led to an increase in the proportion of viable spermatozoa and a decrease in the proportion of late apoptotic/necrotic spermatozoa. No significant change was observed in the proportion of early apoptotic spermatozoa, a finding that supports the concept of abortive apoptosis – namely, that a portion of ejaculated spermatozoa physiologically resides in the initial stages of apoptosis [34,35].

In the case of asymptomatic urogenital infections caused by *Ureaplasma urealyticum*, *Mycoplasma hominis*, *Mycoplasma genitalium*, and *Chlamydia trachomatis*, doxycycline 100 mg twice daily for 7-10 days remains the drug of first choice showing high efficacy and low resistance rates [20], alternatively followed by azithromycin 1 g as a single dose for *Ureaplasma urealyticum* and *Chlamydia trachomatis* [36], or moxifloxacin 400 mg once daily for 7 days, however, emerging resistance is a concern in the latter one [22].

Beyond the clinical management of infection, it is important to emphasize that, as these infections are transmitted through sexual intercourse, the mere fact of being diagnosed with a sexually transmitted infection may stigmatize the couple, with potentially serious consequences for their relationship if proper counselling is not provided – particularly as these are often couples already under considerable psychological strain from long-standing infertility. This implies that, due to the asymptomatic nature of the disease, it is impossible to determine when and from whom someone became infected, and that the presence of the infection does not necessarily mean infidelity in a couple. Last but not least, it is also worth mentioning that non-sexual transmission of the bacteria in question cannot be ruled out.

The high prevalence of some of these bacteria – e.g., *Ureaplasmas*, isolated from cervical or vaginal specimens in up to 40–80% of sexually active asymptomatic women, and from urethral or urine specimens in 20–60% of sexually active men [37] – raises the question of whether this represents a true pathogen or rather a harmless component of urogenital mucosal flora not requiring treatment. Against the latter interpretation, however, stands the finding that *Ureaplasma urealyticum* was detected significantly more frequently in women with preterm labour or preterm premature rupture of membranes (43.9%) than in a comparison group undergoing caesarean delivery for other indications (2.7%) [38]. It is thus assumed that impaired sperm quality, or infertility in general, is not primarily attributable to the mere long-term presence of the bacteria themselves, but rather to a chronic inflammatory immune response, which results in the formation of ROS that damage cell membranes through lipid peroxidation [39].

Not only the pathogen itself, but most importantly the accompanying inflammation and resulting post-infective changes, comprising scarring leading to strictures of the genital tract, or pelvic inflammatory disease, are the underlying mechanisms of damage [13,40]. There are conflicting data, whether similar negative impact on fertility could be established also in couples infected with Human papilomavirus (HPV) [41–45] or *Trichomonas vaginalis* [3].

It is worth mentioning that the causative bacteria may no longer be present at the time of diagnosis – i.e., post-infectious inflammation – or the inflammation may be primarily non-infectious in origin (e.g., autoimmune).

The presence of non-infectious inflammation has traditionally been suspected based on the detection of leukocytes in the ejaculate – leukocytospermia – in combination with a negative microbiological examination. In such cases, conventional semen analysis describes the presence of so-called round cells, which, in addition to leukocytes, may represent spermatids, i.e., developmental stages of sperm [46]. Leukocytes can be distinguished from immature germ cells by peroxidase staining or, cytometrically, by labelling with antibodies specific to CD45.

Flow cytometry also allows differentiation of individual leukocyte subpopulations, namely neutrophils from macrophages, whose ratio can vary significantly among individuals [47]. Given that macrophages are more potent producers of inflammatory cytokines, they are believed to exert a more pronounced negative effect on spermatogenesis. This variable ratio of leukocyte subpopulations may at least partly explain the heterogeneity of published data on the effect of sterile leukocytospermia on ejaculate quality [48].

Leukocytospermia is defined as the presence of more than 1 million peroxidase-positive leukocytes per mL of ejaculate; however, this threshold is arbitrary and has insufficient sensitivity (51–56%), particularly for diagnosing inflammation localized to the epididymis (which contributes only approximately 1% of ejaculate volume) [46]. For this reason, some authors have proposed lower thresholds for leukocytospermia, as low as 0.315 million/mL [49], or even 0.2 million/mL [39,50]. When sterile leukocytospermia is detected, determination of pro-inflammatory cytokines (e.g., tumor necrosis factor alpha, interleukin-6) or chemotactic factors (e.g., interleukin-8) in seminal plasma by enzyme-linked immunosorbent assay (ELISA) may be a useful addition to the diagnostic algorithm [51,52]. If an ongoing non-infectious inflammatory reaction is confirmed, treatment with nonsteroidal anti-inflammatory drugs or the mast cell stabilizer ketotifen – targeting mast cells as a possible source of proinflammatory cytokines in the ejaculate – may be recommended [2,48].

Current evidence suggests that seminal cytokine profiling is more sensitive for detecting inflammation, with interleukin-8 yielding highest sensitivity and specificity [53], than the traditionally used leukocyte count in ejaculate [49]. As our study did not include cytokine profiling or leukocyte subpopulation analysis, we cannot exclude that residual, non-infectious inflammation contributed to the heterogeneity of treatment response observed in our cohort – including the attenuated effect seen beyond 6 months after treatment completion. Incorporating cytokine-based markers into future studies of antibiotic treatment for asymptomatic urogenital infection could help clarify whether persistent inflammation, rather than reinfection alone, underlies this pattern.

## Limitations

This study has several limitations. First, this was a single-centre retrospective study without a control group, rather than a multi-centre prospective randomized trial. While the paired pre- and post-treatment semen analysis in the same individual provides a relatively robust within-subject comparison, the absence of a control group limits our ability to draw causal conclusions regarding the effect of antibiotic treatment on the female partner’s likelihood of conception, as confounding factors on the female side – including age, gynaecological findings, and oocyte quality, the strongest known predictors of couple infertility – could not be accounted for.

Second, as outlined in the Introduction, the physiology of human reproduction itself imposes inherent limitations on the interpretation of semen analysis data. Conventional semen parameters can fluctuate considerably over time even in fertile men. Given that sperm generation time is nearly three months, men are inevitably exposed during this period to a range of poorly defined and uncontrolled factors beyond strictly defined study conditions. Furthermore, infertility represents merely a phenotype arising from a broad spectrum of underlying causes, and no identifiable cause is found in 30-40% of men despite thorough andrological work-up, resulting in a diagnosis of idiopathic infertility. Abnormalities in conventional semen parameters are non-specific, meaning that a given finding cannot reliably indicate the underlying cause of infertility; conversely, even when a specific cause is identified, it is not certain that it is truly causally responsible for reduced fertility potential. Moreover, conventional semen analysis is fundamentally a morphological assessment, and its parameters do not necessarily correlate with the actual probability of conception.

Third, partner concordance analysis was based on the subset of couples with complete bilateral testing (n=244).

Fourth, total sperm motility was recorded systematically only from July 2020 onward and was therefore analysed in a smaller subset of the cohort (130 of 265 pairs) than the remaining parameters. The assessment of sperm morphology was not consistent throughout the study period, precluding its inclusion in the analysis.

Fifth, no microbiological test-of-cure was performed following antibiotic treatment, and eradication of the causative organisms was therefore not formally confirmed.

Sixth, due to the retrospective design, women were not uniformly exposed to assisted reproductive techniques (ART) at the same time interval following completion of antibiotic treatment, as some couples opted for repeated intrauterine insemination first, while others proceeded early to IVF with intracytoplasmic sperm injection (ICSI). Pregnancy was confirmed either by the treating gynaecologist at our institution or by the woman’s registering gynaecologist and subsequently ascertained by the study team through patient self-report, which may be subject to reporting bias.

## 5. Conclusions

Antibiotic treatment of asymptomatic urogenital infections in infertile couples was associated with significant improvements in conventional semen parameters, including sperm concentration, total sperm count, and both total and progressive motility, as well as a reduction in the proportion of late apoptotic / necrotic sperm and calculated sperm DNA fragmentation. These improvements were most pronounced within the first three months after treatment and appeared to diminish beyond six months. Nearly half of the couples achieved pregnancy within 12 months of treatment completion, most frequently through ART. Given the retrospective, single-centre design without a control group, these findings cannot establish a causal relationship between antibiotic treatment and improved fertility outcomes. Nonetheless, they support the pragmatic approach of treating both partners of an infertile couple upon detection of asymptomatic urogenital infection in either partner, and provide a rationale for future prospective, controlled studies.

## Author contributions

(1) Conception and design: Z.K., V.V., A.L., J.N.; (2) Administrative support: J.N., Z.K.; (3) Provision of study material or patients: J.N., Z.K., V.V., A.L.; (4) Collection and assembly of data: Z.K., J.N.; (5) Data analysis and interpretation: M.D., V.V., Z.K., J.N.; (6) Manuscript writing: All authors; (7) Final approval of manuscript: All authors.

## Data Availability

All data produced in the present study are available upon reasonable request to the authors

## Acknowledgements

Not applicable.

## Footnote

### Funding

None.

### Ethical Statement

The authors are accountable for all aspects of the work in ensuring that questions related to the accuracy or integrity of any part of the work are appropriately investigated and resolved. The study was conducted in accordance with the Declaration of Helsinki (as revised in 2013). The requirement for ethics committee approval was waived by the Ethics Committee of IVF Clinic GENNET, Prague, Czech Republic, given the retrospective, anonymized analysis of data obtained during routine clinical care. All participants provided written informed consent for the use of their anonymized clinical data for statistical analysis and publication.

## List of abbreviations

ANOVA: analysis of variance
ART: assisted reproductive technologies
BF_10_: Bayes factor
CI: confidence interval
CLES: common language effect size
DGGG: Deutsche Gesellschaft für Gynäkologie und Geburtshilfe (German gynaecological and obstetric society)
EAU: European Association of Urology
EPS: expressed prostatic secretions
FACS-APO: flow cytometry sperm apoptosis test
FITC: fluorescein isothiocyanate
ICSI: intracytoplasmic sperm injection
IQR: interquartile range
IUI: intrauterine insemination
IVF: in vitro fertilization
MAGIs: male accessory gland infections
NPV: negative predictive value
OEGGG: Österreichische Gesellschaft für Gynäkologie und Geburtshilfe (Austrian gynaecological and obstetric society)
PI: propidium iodide
RBC: rank-biserial correlation
ROS: reactive oxygen species
SD: standard deviation
SGGG: Schweizerische Gesellschaft für Gynäkologie und Geburtshilfe (Swiss gynaecological and obstetric society)
WHO: World Health Organization

